# *PCSK9* genetic variants, carotid atherosclerosis and vascular remodelling

**DOI:** 10.1101/2025.02.25.25322905

**Authors:** Daniela Coggi, Joey Ward, Bruna Gigante, Mauro Amato, Donald M. Lyall, Beatrice Frigerio, Alessio Ravani, Daniela Sansaro, Nicola Ferri, Maria Giovanna Lupo, Chiara Macchi, Massimiliano Ruscica, Fabrizio Veglia, Nicolò Capra, Antonio Gallo, Matteo Pirro, Kai Savonen, Douwe J. Mulder, Roberta Baetta, Elena Tremoli, Jill P. Pell, Paul Welsh, Naveed Sattar, Damiano Baldassarre, Rona J. Strawbridge, the IMPROVE study group

## Abstract

**Background and aims:** Circulating PCSK9 is a crucial regulator of cholesterol metabolism. Loss-of-function variants in PCSK9 are associated with lower levels of circulating low-density lipoprotein cholesterol (LDL-C) and reduced cardiovascular disease (CVD) risk, while gain-of-function variants correlate with elevated LDL-C concentrations and increased CVD risk. This study investigated whether genetically determined LDL-C levels, proxied by four PCSK9 genetic variants, influence common carotid artery atherosclerosis.

**Methods:** The analysis included 3,040 European participants (mean age 64.2 ± 5.4 years; 45.8% men) at high cardiovascular risk from the IMPROVE study, alongside 49,088 individuals of white British ancestry (mean age 55.2 ± 7.6 years; 47.9% men) from the UK Biobank (UKB). Ultrasonographic measurements of common carotid intima-media thickness (CC-IMT_mean_, CC-IMT_max_, CC-IMT_mean-max_) were obtained. Four lipid-level affecting genetic variants in the *PCSK9* locus were selected for analysis, both individually and in a standardized polygenic risk score (PRS), to assess their effects on LDL-C and PCSK9 levels in the IMPROVE cohort and on ultrasonographic measures in both IMPROVE and UKB.

**Results:** In the IMPROVE cohort, *PCSK9* variants (rs11206510, rs2479409, rs11591147, rs11583680) exhibited expected effect directions, albeit not all statistically significant, on LDL-C and PCSK9 levels. The PRS was negatively correlated with CC-IMT_mean_, CC-IMT_max_, and CC-IMT_mean-max_ among women in IMPROVE, and among men and overall in UKB (all P < 0.05). Effect sizes were comparable between cohorts.

**Conclusions:** Genetic variants in the *PCSK9* locus influence LDL-C levels and CC-IMT, in keeping with proven benefits of PCSK9 inhibitors on atherosclerotic cardiovascular events.

## Introduction

Intima-media thickness (IMT) of extracranial carotid arteries, measured non-invasively using B-mode ultrasound, is an imaging measure of arterial wall thickness often used as a surrogate marker of clinical ^1^ and subclinical atherosclerosis ^2^. IMT is directly associated with all traditional cardiovascular risk factors, such as hypercholesterolemia ^3^, age^3^, blood pressure ^4^, diabetes ^5^ , insulin resistance ^6^ and cigarettes smoking ^7^, as well as with the prevalent^8^and incident ^8^ vascular events. More recently, some^9, 10^, but not all ^11, 12^, studies demonstrated that circulating levels of proprotein convertase subtilisin/kexin type 9 (PCSK9) are also associated with carotid IMT. PCSK9 (encoded by the *PCSK9* gene) is a serine protease mainly synthesized and secreted by the liver that plays a role not only in atherosclerosis ^13^, but also in the development of cardiovascular diseases (CVD) ^14^.

PCSK9 post-transcriptionally causes the degradation of the low-density-lipoprotein receptor (LDLR) ^15^ and thus indirectly increases plasma low-density-lipoprotein cholesterol (LDL-C) levels ^15^. Since 2003, many *PCSK9* genetic variants with “gain-of-function” (GOF) ^14^ or “loss-of-function” (LOF) ^14^ effects have been described. In particular, GOF variants were consistently associated with higher LDL-C levels ^14^ through reduced LDLR levels, resulting in a higher risk of CVD ^14^ or cerebrovascular diseases ^16^. Conversely, LOF variants were consistently associated with lower LDL-C concentrations^14, 17–21^ through increased LDLR levels, resulting in a lower risk of CVD^14, 17, 19, 21, 22^. PCSK9 inhibitors such as evolocumab and alirocumab are efficacious in reducing cardiovascular events in patients at high risk ^23^ and are recommended for patients with uncontrolled familial hypercholesterolaemia ^24^. While many studies have evaluated the relationship between *PCSK9* variants and CVD or cerebrovascular diseases, to our knowledge few studies have evaluated the relationship between *PCSK9* variants and subclinical atherosclerosis. Only one study has investigated the effect of the GOF E670G (rs505151) and I474V (rs562556) variants on carotid IMT ^25^.

Similarly, only one study has investigated the effect of the LOF variants Y142X (rs67608943), C679X (rs28362286) ^19^ and two studies have investigated the effect of R46L (rs11591147) variant ^17, 19^.

In this study, we extend previous work by investigating the impact of four genetic variants known to influence LDL-C levels ^26^, individually and combined in a polygenic risk score, on carotid IMT measures in a high CVD-risk cohort (IMPROVE) and a large general population cohort (UK Biobank (UKB)). As there are known differences in CVD by sex ^27^, and as lipid-lowering medication might influence IMT (through PCSK9 dependent or independent mechanisms), the possibility of sex-specific or treatment-dependent interactions was also evaluated.

## Material and methods

### Selection of PCSK9 variants

The selection of four variants in the *PCSK9* locus was based on the Schmidt et al.^26^ definition, namely: (1) a robust association with LDL-C (as observed by the Global Lipids Genetics Consortium^28^), (2) a low pairwise linkage disequilibrium (LD) (r^2^≤0.30) with other variants in that region (using 1000 Genomes CEU data); and (3) a combined annotation dependent depletion (CADD) score^29^ that evaluates potential functionality. Specifically, the four variants were: rs11206510 (build 37, chromosome 1:55505668, C allele lipid lowering), rs2479409 (1:55504650, G allele lipid increasing), rs11591147 (i.e., R46L, 1:55505647, T allele lipid lowering) and rs11583680 (1:55505668, T allele lipid lowering).

### Study cohorts and phenotyping

The IMPROVE study design, eligibility criteria, aims, and baseline evaluation have already been reported in detail ^30^. Briefly, a total of 3,711 subjects (age range 54‒79 years), defined as high CVD risk based on the presence of at least three traditional vascular risk factors (e.g. hypercholesterolemia, hypertension, diabetes) but free from CVD and cerebrovascular diseases, were recruited in seven centres across five European countries, i.e. Finland (Kuopio, two centres), Sweden (Stockholm), the Netherlands (Groningen), France (Paris), and Italy (Milan and Perugia) between January 2004 and June 2005. In 2023, the Institute of Public Health and Clinical Nutrition at the University of Eastern Finland in Kuopio revoked data usage permission for participants recruited to this centre. As a result, this study included the 3,040 participants recruited from the remaining six centres.

During the baseline visit, participants completed a medical history, medication and lifestyle questionnaire, carotid ultrasound examination and blood sampling for standard biochemical tests and genotyping. Details of lipid measurements and carotid ultrasound examination have been previously published ^30^ and are summarised in the Supplemental Material. For this study, three indices of the common carotid (CC) arterial wall damage (CC-IMT_mean_, CC-IMT_max_ and CC-IMT_mean-max_) were used, as these were deemed most comparable to the measurements conducted by UKB. Circulating PCSK9 levels were measured using commercial ELISA kits (R&D Systems, MN) able to recognize free and LDLR-bound PCSK9 ^31^ (Supplemental Material).

The IMPROVE study complies with the rules of Good Clinical Practice and with the ethical principles established in the Declaration of Helsinki, and was approved by 6 independent ethics committees, i.e.: the Regional Ethics Review Board at Karolinska Institutet, Stockholm, Sweden (approval ID: Dnr 2003/03-115, 17 February 2003); the Institutional Review Board of the Health Department of the Hospital “Ospedale Niguarda Ca’ Granda”, Milan, Italy (approval ID: 2042/03, 3 June 2003); the Medical Ethics Review Committee, Academic Hospital Groningen, Groningen, the Netherlands (approval ID: METc 2003/054, 12 May 2003); the Ethics Committee of the Umbrian Health Authorities, Perugia, Italy (approval ID: N 2725/03/A, 6 February 2003); the Consultative Committee for the Protection of Persons in Biomedical Research (CCPPRB) at Hôspital Pitie Salpétrière, Paris, France (approval ID: CCP61-03, 2003); the Research Ethics Committee of the University of Kuopio and Kuopio University Hospital, Kuopio, Finland (approval ID: 140/2002, 10 October 2002). All patients provided informed consent twice; once for general participation in the study and once for genotyping.

The UKB cohort has also been described previously ^32^. In short, UKB included over 500,000 participants (age range 40–69 at baseline) recruited from the general population who attended one of 22 centres across the UK between 2006 and 2010. A self-completed baseline questionnaire provided information on personal, and family medical history, medication, lifestyle and blood samples were taken for standard biochemical assays and genotyping. A subset of individuals was invited to participate in a follow-up imaging visit (4-8 years after baseline), which included carotid ultrasound examination. For this study, only 49,088 unrelated self-reported white British ancestry individuals with CC ultrasonographic variables measures were included. Details of the carotid ultrasound examination have been published previously ^33^ and are summarised in the Supplemental Material.

All participants in UKB provided informed electronic consent and UKB was granted ethical approval by the North-West Multicentre Research Ethics Committee on the 29^th^ of June 2021 (Ref 21/NW/0157). This study was conducted under project #71392, (P.I. Strawbridge).

### Genetic data

IMPROVE subjects were genotyped using both Illumina Cardio-Metabo 200k ^34^ and Immunochip 200k ^35^ array platforms. Standard quality control was conducted on the individual chips as well as the combined dataset, including: exclusion of samples with cryptic relatedness, low call rate (<95%) or ambiguous sex and exclusion of variants for low call rate (<95%), failing Hardy-Weinberg Equilibrium (p<1x10^-5^) or low call rate (minor allele frequency (MAF) >1%). Multi-dimensional scaling (MDS) components 1, 2, and 3 were calculated in PLINK ^36^ to enable adjustment for population structure.

UKB participants were genotyped using either the Affymetrix BiLEVE Axion or Affymetrix UKB Axion array and imputed to 1000 Genomes, UK10K and Haplotype Reference Consortium (March 2018 release) reference panels ^37^. Standard genetic quality control (consistent with that described above), pre- and post-imputation, was conducted centrally by UKB. Genetic principal components were also computed by the central UKB team. Eight principal genetic components (PGC1-8) were used to enable adjustment for population structure. Only unrelated individuals of self-reported white British ancestry were included in the analyses.

The MAF and Hardy-Weinberg equilibrium *p* of variants in IMPROVE and UKB are shown in S Table 1. The LD between the variants in IMPROVE and UKB was investigated with r^2^=0.22 and 0.28 respectively between variants rs11206510 and rs11583680. The LD between other variant pairs indicated independent effects for both IMPROVE (r^2^≤0.07) (S Figure 1, Panel A) and UKB (r^2^≤0.08) (S Figure 1, Panel B).

**Figure 1.**
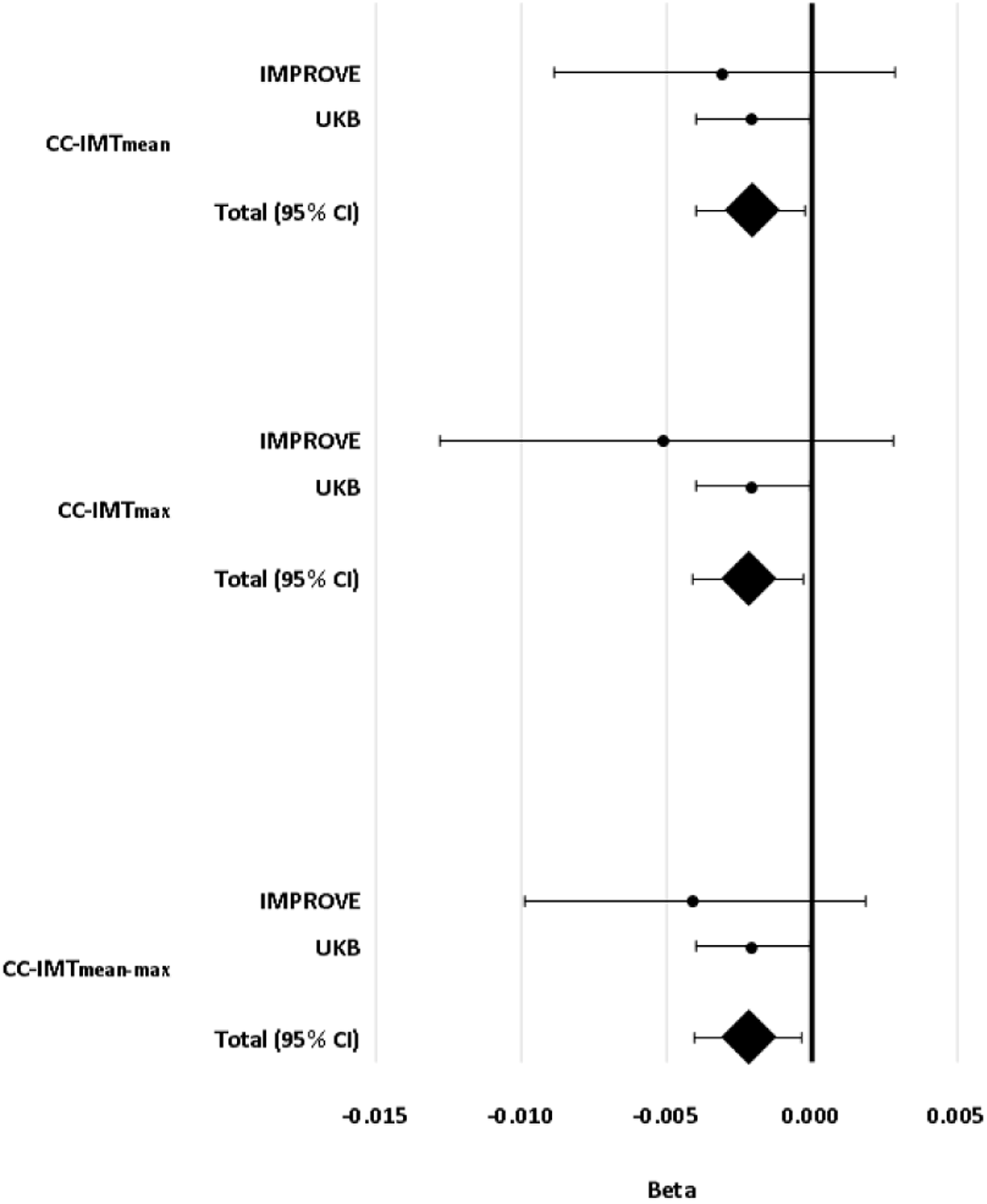
Individual betas (IMPROVE and UKB) and summarize beta (95% CI) of PRS on CC-IMT_mean_, CC- IMT_max_ and CC-IMT_mean-max_. Beta adjusted for sex, age, population structure (MDS1-3) and use of lipid-lowering medication in IMPROVE. Beta adjusted for sex, age, population structure (PGC1-8), genotyping chip and lipid-lowering medication in UKB. CC-IMT_mean_, average of means of intima-media thickness in left and right common carotid arteries; CC-IMT_max_, the highest value of maximum intima-media thickness in left and right common carotid arteries; CC-IMT_mean-max_, mean of maximum intima-media thickness in left and right common carotid arteries. *P* < 0.05 were considered statistically significant.

**Table 1.** Baseline characteristics of the IMPROVE and UKB studies cohorts.

|  | IMPROVE |  |  | UKB |  |  |
| --- | --- | --- | --- | --- | --- | --- |
|  | Men | Women | All | Men | Women | All |
| N (%) | 1,393 (45.8) | 1,647 (54.2) | 3,040 | 23,506 (47.9) | 25,582 (52.1) | 49,088 |
| Age, years | 64.0 ± 5.3 | 64.4 ± 5.5 | 64.2 ± 5.4 | 55.9 ± 7.7 | 54.9 ± 7.7 | 55.2 ± 7.6 |
| BMI, kg/m<sup>2</sup> | 27.5 ± 3.7 | 26.9 ± 4.7 | 27.2 ± 4.3 | 27.1 ± 3.7 | 26.0 ± 4.5 | 26.5 ± 4.2 |
| Waist/hip ratio | 0.97 ± 0.07 | 0.87 ± 0.07 | 0.92 ± 0.09 | 0.92 ± 0.06 | 0.80 ± 0.07 | 0.86 ± 0.09 |
| LDL-C, mmol/L | 3.43 ± 0.94 | 3.75 ± 1.05 | 3.61 ± 1.01 | na | na | na |
| HDL-C, mmol/L | 1.12 ± 0.30 | 1.37 ± 0.37 | 1.26 ± 0.36 | na | na | na |
| Triglycerides, mmol/L | 1.45 (1.02; 2.11) | 1.27 (0.92; 1.81) | 1.34 (0.96; 1.95) | na | na | na |
| PCSK9, ng/mL | 288 (227; 358) | 326 (266; 395) | 307 (247; 381) | na | na | na |
| Lipid-lowering medication, n (%) | 676 (48.7) | 822 (49.9) | 1,498 (49.4) | 3,971 (17.2) | 1,801 (7.2) | 5,772 (12.0) |
| CC-IMT<sub>mean</sub>, mm | 0.75 (0.67; 0.85) | 0.71 (0.65; 0.78) | 0.73 (0.66; 0.81) | 0.70 (0.61; 0.80) | 0.65 (0.59; 0.73) | 0.67 (0.60; 0.76) |
| CC-IMT<sub>max</sub>, mm | 1.16 (1.02; 1.45) | 1.06 (0.97; 1.21) | 1.10 (0.98; 1.31) | 0.92 (0.81; 1.08) | 0.85 (0.77; 1.00) | 0.89 (0.77; 1.04) |
| CC-IMT<sub>mean-max</sub>, mm | 1.05 (0.94; 1.21) | 0.98 (0.91; 1.10) | 1.01 (0.92; 1.14) | 0.81 (0.71; 0.93) | 0.76 (0.67; 0.85) | 0.78 (0.69; 0.89) |
| P.H. of stroke, n (%) | na | na | na | 223 (1.27) | 126 (0.59) | 349 (0.90) |
| P.H. of IHD, n (%) | na | na | na | 902 (4.96) | 248 (1.16) | 1,150 (2.90) |
| PRS | 1.90 ± 1.26 | 1.98 ± 1.26 | 1.94 ± 1.26 | 2.00 ± 1.33 | 1.99 ± 1.33 | 2.00 ± 1.33 |

Where: CC-IMT<sub>mean</sub>, average of means of intima-media thickness in left and right common carotid arteries; CC-IMT<sub>max</sub>, the highest value of maximum intima-media thickness in left and right common carotid arteries; CC-IMT<sub>mean-max</sub>, mean of maximum intima-media thickness in left and right common carotid arteries; P.H., personal history; IHD, ischemic heart disease; PRS, polygenic risk score. Binary variables are presented as n (%) and continuous variables are presented as mean ± SD or median (25%; 75%).

### Power calculations

For power calculations for individual variant analyses, we used a conservative estimate of variance explained for a model including age, sex and population structure of 0.001 (based on previous evidence of genetic effect sizes on CC-IMT ^33^), which meant IMPROVE had 40% power to detect an effect, whilst UKB had >99% power.

### Statistical analyses

All continuous phenotypes were evaluated for normality. Triglycerides, PCSK9 and CC-IMT ultrasonographic variables were log normalized prior to analyses. Analyses of individual genetic variants were conducted using linear regression in PLINK 1.07, assuming an additive genetic model.

An unweighted lipid-lowering polygenic risk score (PRS) was constructed by summing the number of LDL-C lowering alleles (0, 1 and 2) of each variant and was used to assess the combined effect of the four variants. Only individuals with complete genotyping were included in PRS analyses. The PRS was standardised and analyses were conducted using general linear models (GLM) in SAS v. 9.4 (SAS Institute Inc., Cary, NC, USA) for IMPROVE and STATA (STATA Corp) for UKB.

In the IMPROVE study, we assessed whether associations were observed between individual variants and lipids and PCSK9. Here, *p*< 0.0125 was considered significant (Bonferroni correction for four comparisons). Subsequently, we investigated the impact of individual variants on CC-IMT_mean_, CC-IMT_max_ and CC-IMT_mean- max_, considering *p*< 0.0125 as significant. Finally, the PRS was tested for associations with CC-IMT_mean_, CC- IMT_max_ and CC-IMT_mean-max_. Here *p*< 0.05 was considered significant. Investigating the associations between individual variants and PCSK9, three models were considered: Model 1, adjusted for sex, age and population structure (i.e., MDS1-3); Model 2, as model 1 plus use of lipid-lowering medication; Model 3, as model 1 plus independent determinants of PCSK9 (use of statins and/or fibrates, total cholesterol, high-density-lipoprotein cholesterol (HDL-C), uric acid, personal history of hypertriglyceridemia and/or hypercholesterolemia, hip circumference, consumption of fish and/or wine, pack-years, family history of diabetes)^31^. Sensitivity analyses were also performed, stratified by sex. Individual variants were also assessed for effects on lipid levels adjusting for Model 2 in the whole group, in men and in women.

The effects on CC-IMT ultrasonographic variables, using Model 2, was tested by considering both the individual variants and the PRS. Sensitivity analyses were also performed, stratified either by sex or by lipid-lowering treatment. A general linear model (GLM) was used to adjust for confounding factors (Model 2) and to evaluate whether (1) PRS was associated with PCSK9 levels after stratification by sex or by use of lipid-lowering medication (*p*< 0.05 as significant); (2) PRS affected PCSK9 levels by interacting with sex or use of lipid-lowering medication (*p*< 0.05 as significant). Additionally, it was employed to estimate adjusted geometric means for PCSK9 levels by accounting for Age and MDS1-3, across each combination of stratification factor values (Women/Men or Treated/Untreated) and PRS.

In UKB, individual variants (*p*< 0.0125 as significant) and the PRS (*P* < 0.05 as significant) were tested for association with CC-IMT_mean_, CC-IMT_max_ and CC-IMT_mean-max_, adjusting for age, sex, genotyping chip, population structure (i.e., PGC1-8) and use of lipid-lowering medication. Again, sensitivity testing included stratification by sex or by use of lipid-lowering medication, and the interaction (*P* < 0.05 as significant) between PRS and sex or between PRS and use of lipid-lowering medication was tested.

To assess whether the very different sample sizes between IMPROVE and UKB might have influenced the results, the estimated beta values of the relationships between PRS and CC ultrasonographic variables from both studies were also analysed with a fixed-effects meta-analysis and visualised in a forest plot.

Heterogeneity was assessed using the Q-test and the I^2^ statistic. *p*< 0.05 were considered significant.

## Results

Demographic features of IMPROVE and UKB participants are presented in Table 1. Men and women were well represented in both studies. Subjects included in the IMPROVE study were about 10 years older than participants of the UKB study. IMPROVE is a high CVD-risk cohort, thus the higher values of CC-IMT_mean_, CC- IMT_max_ and CC-IMT_mean-max_ observed in IMPROVE compared to the general population UKB are unsurprising, despite the more frequent use of lipid-lowering medication in IMPROVE compared to UKB (49.4% vs. 12.0%, respectively).

### Associations of PCSK9 variants with lipids and PCSK9 levels in IMPROVE

The effects rs11206510-C, rs11591147-T, rs11583680-T (all lipid lowering alleles), and rs2479409-G (lipid increasing allele) on lipids levels and PCSK9 in the IMPROVE study are shown in Table 2. Of these variants, only rs11591147-T (i.e., R46L) was significantly associated with LDL-C levels with the effect directions (i.e., beta <0) in line with expectation (Table 2). No associations were observed between these variants and HDL-C or triglycerides. All four variants were associated with PCSK9 levels, with the expected directions of effect: rs11206510-C, rs11591147-T, and rs11583680-T were associated with lower PCSK9 levels and rs2479409-G was associated with elevated PCSK9 levels. Effect directions were similar in sex-stratified analyses, although not all associations reached statistical significance (*P* < 0.0125).

**Table 2.**
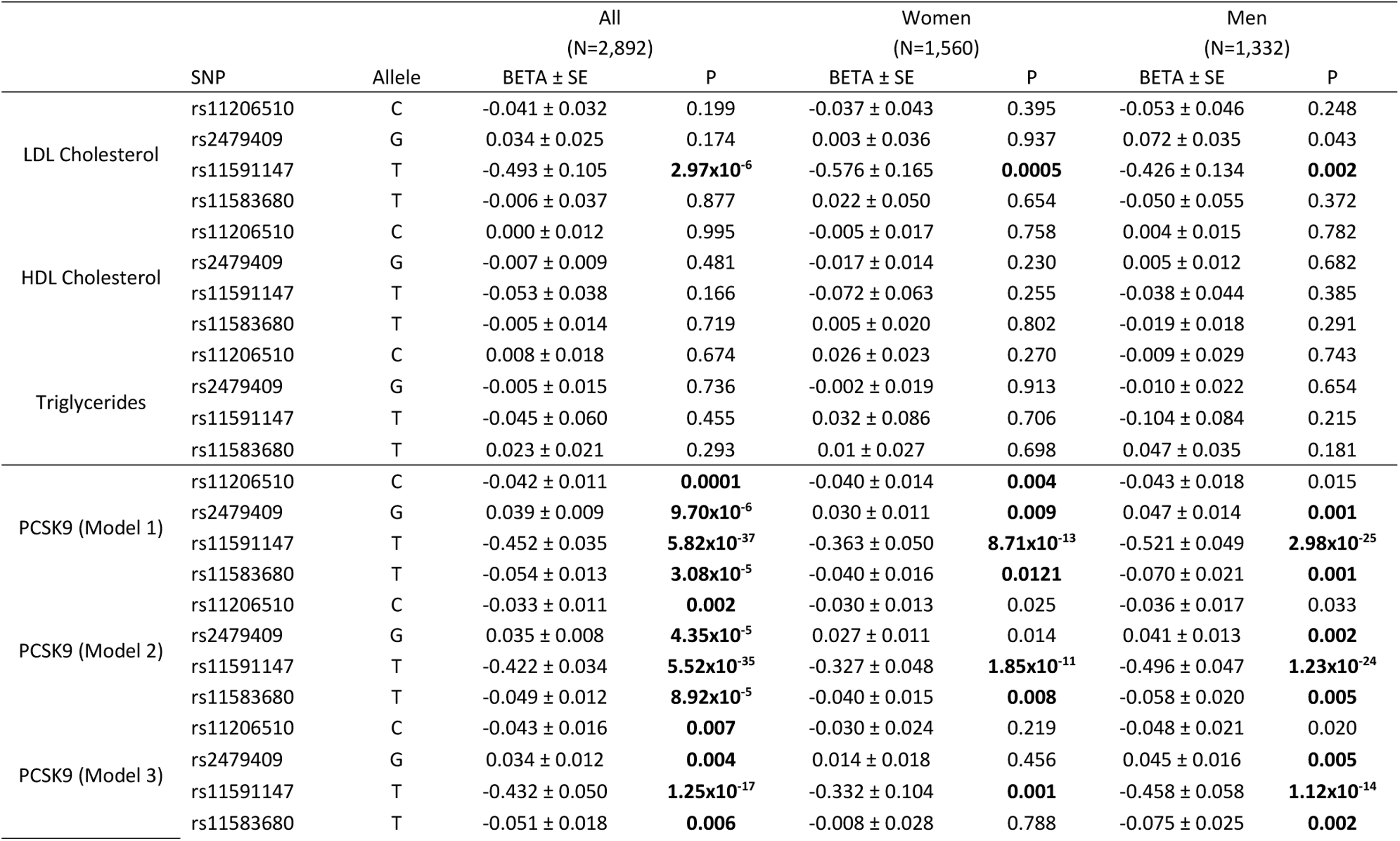
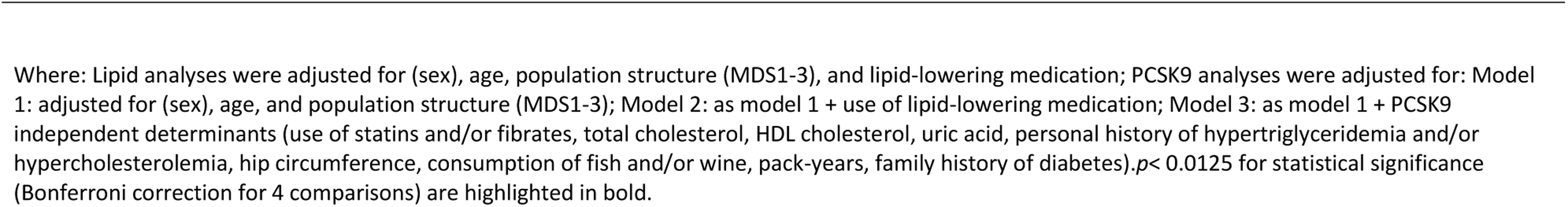
Association between single variants and lipids and PCSK9 measures in the IMPROVE study.

|  | SNP | Allele | All<br>(N=2,892) |  | Women<br>(N=1,560) |  | Men<br>(N=1,332) |  |
| --- | --- | --- | --- | --- | --- | --- | --- | --- |
|  |  |  | BETA ± SE | P | BETA ± SE | P | BETA ± SE | P |
| LDL Cholesterol | rs11206510 | C | -0.041 ± 0.032 | 0.199 | -0.037 ± 0.043 | 0.395 | -0.053 ± 0.046 | 0.248 |
|  | rs2479409 | G | 0.034 ± 0.025 | 0.174 | 0.003 ± 0.036 | 0.937 | 0.072 ± 0.035 | 0.043 |
|  | rs11591147 | T | -0.493 ± 0.105 | **2.97x10<sup>-6</sup>** | -0.576 ± 0.165 | **0.0005** | -0.426 ± 0.134 | **0.002** |
|  | rs11583680 | T | -0.006 ± 0.037 | 0.877 | 0.022 ± 0.050 | 0.654 | -0.050 ± 0.055 | 0.372 |
| HDL Cholesterol | rs11206510 | C | 0.000 ± 0.012 | 0.995 | -0.005 ± 0.017 | 0.758 | 0.004 ± 0.015 | 0.782 |
|  | rs2479409 | G | -0.007 ± 0.009 | 0.481 | -0.017 ± 0.014 | 0.230 | 0.005 ± 0.012 | 0.682 |
|  | rs11591147 | T | -0.053 ± 0.038 | 0.166 | -0.072 ± 0.063 | 0.255 | -0.038 ± 0.044 | 0.385 |
|  | rs11583680 | T | -0.005 ± 0.014 | 0.719 | 0.005 ± 0.020 | 0.802 | -0.019 ± 0.018 | 0.291 |
| Triglycerides | rs11206510 | C | 0.008 ± 0.018 | 0.674 | 0.026 ± 0.023 | 0.270 | -0.009 ± 0.029 | 0.743 |
|  | rs2479409 | G | -0.005 ± 0.015 | 0.736 | -0.002 ± 0.019 | 0.913 | -0.010 ± 0.022 | 0.654 |
|  | rs11591147 | T | -0.045 ± 0.060 | 0.455 | 0.032 ± 0.086 | 0.706 | -0.104 ± 0.084 | 0.215 |
|  | rs11583680 | T | 0.023 ± 0.021 | 0.293 | 0.01 ± 0.027 | 0.698 | 0.047 ± 0.035 | 0.181 |
| PCSK9 (Model 1) | rs11206510 | C | -0.042 ± 0.011 | **0.0001** | -0.040 ± 0.014 | **0.004** | -0.043 ± 0.018 | 0.015 |
|  | rs2479409 | G | 0.039 ± 0.009 | **9.70x10<sup>-6</sup>** | 0.030 ± 0.011 | **0.009** | 0.047 ± 0.014 | **0.001** |
|  | rs11591147 | T | -0.452 ± 0.035 | **5.82x10<sup>-37</sup>** | -0.363 ± 0.050 | **8.71x10<sup>-13</sup>** | -0.521 ± 0.049 | **2.98x10<sup>-25</sup>** |
|  | rs11583680 | T | -0.054 ± 0.013 | **3.08x10<sup>-5</sup>** | -0.040 ± 0.016 | **0.0121** | -0.070 ± 0.021 | **0.001** |
| PCSK9 (Model 2) | rs11206510 | C | -0.033 ± 0.011 | **0.002** | -0.030 ± 0.013 | 0.025 | -0.036 ± 0.017 | 0.033 |
|  | rs2479409 | G | 0.035 ± 0.008 | **4.35x10<sup>-5</sup>** | 0.027 ± 0.011 | 0.014 | 0.041 ± 0.013 | **0.002** |
|  | rs11591147 | T | -0.422 ± 0.034 | **5.52x10<sup>-35</sup>** | -0.327 ± 0.048 | **1.85x10<sup>-11</sup>** | -0.496 ± 0.047 | **1.23x10<sup>-24</sup>** |
|  | rs11583680 | T | -0.049 ± 0.012 | **8.92x10<sup>-5</sup>** | -0.040 ± 0.015 | **0.008** | -0.058 ± 0.020 | **0.005** |
| PCSK9 (Model 3) | rs11206510 | C | -0.043 ± 0.016 | **0.007** | -0.030 ± 0.024 | 0.219 | -0.048 ± 0.021 | 0.020 |
|  | rs2479409 | G | 0.034 ± 0.012 | **0.004** | 0.014 ± 0.018 | 0.456 | 0.045 ± 0.016 | **0.005** |
|  | rs11591147 | T | -0.432 ± 0.050 | **1.25x10<sup>-17</sup>** | -0.332 ± 0.104 | **0.001** | -0.458 ± 0.058 | **1.12x10<sup>-14</sup>** |
|  | rs11583680 | T | -0.051 ± 0.018 | **0.006** | -0.008 ± 0.028 | 0.788 | -0.075 ± 0.025 | **0.002** |

Where: Lipid analyses were adjusted for (sex), age, population structure (MDS1-3), and lipid-lowering medication; PCSK9 analyses were adjusted for: Model 1: adjusted for (sex), age, and population structure (MDS1-3); Model 2: as model 1 + use of lipid-lowering medication; Model 3: as model 1 + PCSK9 independent determinants (use of statins and/or fibrates, total cholesterol, HDL cholesterol, uric acid, personal history of hypertriglyceridemia and/or hypercholesterolemia, hip circumference, consumption of fish and/or wine, pack-years, family history of diabetes). $p < 0.0125$ for statistical significance (Bonferroni correction for 4 comparisons) are highlighted in bold.

### Associations of PCSK9 variants with CC-IMT variables in IMPROVE

In IMPROVE, no significant associations were observed between individual *PCSK9* variants and CC-IMT_mean_, CC-IMT_max_ and CC-IMT_mean-max_ in sex-combined or sex-stratified analyses (S Table 2). However, effect directions were consistent with expectations (i.e., beta >0 for rs2479409-G and beta <0 for rs11206510-C, rs11591147-T or rs11583680-T) for CC-IMT_max_ and CC-IMT_mean-max_ in sex-combined analyses, and for CC- IMT_max_ in women.

In IMPROVE, no significant associations were observed when analyses were stratified by lipid-lowering treatment (S Table 3). In untreated subjects, the effect direction was in line with that expected for all variants. In subjects treated with lipid lowering drugs, instead, the effect direction for rs2479409-G was in line with that expected (i.e. beta >0) for all CC-IMT variables, whereas the effect direction for rs11206510-C has the expected direction (beta <0) for CC-IMT_mean_ and CC-IMT_mean-max_ only.

**Table 3:**
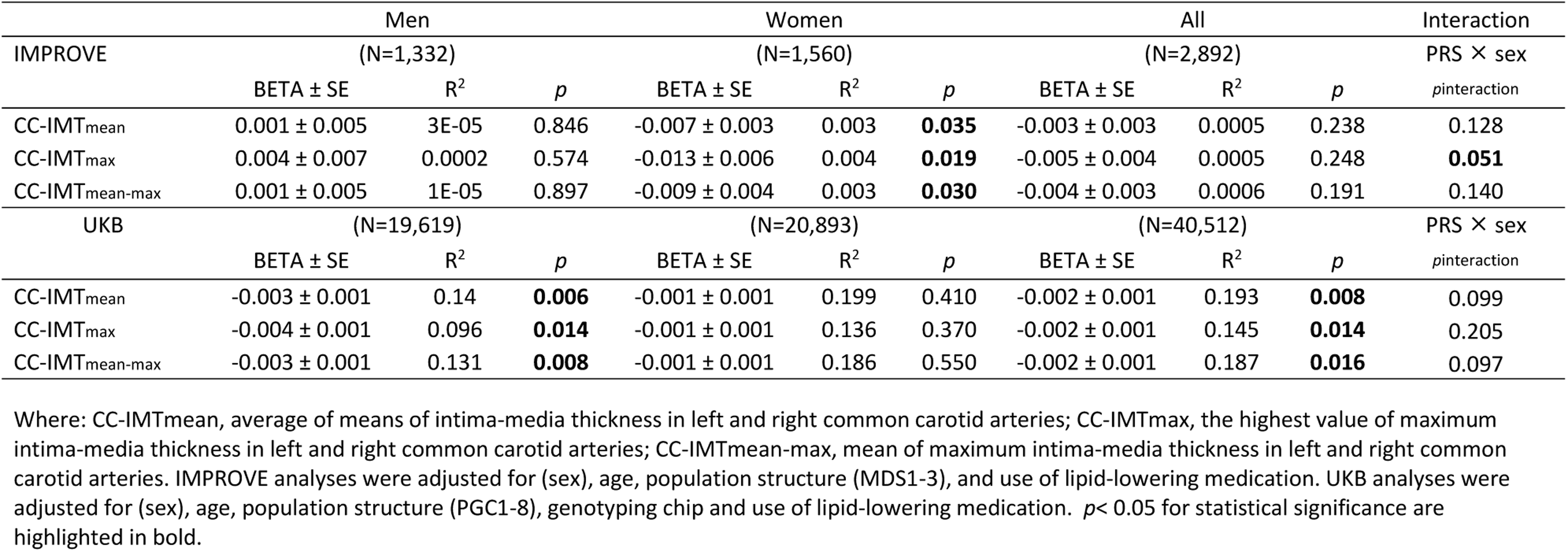
Associations of the lipid-lowering PRS with CC-IMT variables in IMPROVE and UKB.

|  | Men |  |  | Women |  |  | All |  |  | Interaction |
| --- | --- | --- | --- | --- | --- | --- | --- | --- | --- | --- |
| IMPROVE | (N=1,332) |  |  | (N=1,560) |  |  | (N=2,892) |  |  | PRS × sex |
|  | BETA ± SE | R<sup>2</sup> | p | BETA ± SE | R<sup>2</sup> | p | BETA ± SE | R<sup>2</sup> | p | p<sub>interaction</sub> |
| CC-IMT<sub>mean</sub> | 0.001 ± 0.005 | 3E-05 | 0.846 | -0.007 ± 0.003 | 0.003 | **0.035** | -0.003 ± 0.003 | 0.0005 | 0.238 | 0.128 |
| CC-IMT<sub>max</sub> | 0.004 ± 0.007 | 0.0002 | 0.574 | -0.013 ± 0.006 | 0.004 | **0.019** | -0.005 ± 0.004 | 0.0005 | 0.248 | **0.051** |
| CC-IMT<sub>mean-max</sub> | 0.001 ± 0.005 | 1E-05 | 0.897 | -0.009 ± 0.004 | 0.003 | **0.030** | -0.004 ± 0.003 | 0.0006 | 0.191 | 0.140 |
| UKB | (N=19,619) |  |  | (N=20,893) |  |  | (N=40,512) |  |  | PRS × sex |
|  | BETA ± SE | R<sup>2</sup> | p | BETA ± SE | R<sup>2</sup> | p | BETA ± SE | R<sup>2</sup> | p | p<sub>interaction</sub> |
| CC-IMT<sub>mean</sub> | -0.003 ± 0.001 | 0.14 | **0.006** | -0.001 ± 0.001 | 0.199 | 0.410 | -0.002 ± 0.001 | 0.193 | **0.008** | 0.099 |
| CC-IMT<sub>max</sub> | -0.004 ± 0.001 | 0.096 | **0.014** | -0.001 ± 0.001 | 0.136 | 0.370 | -0.002 ± 0.001 | 0.145 | **0.014** | 0.205 |
| CC-IMT<sub>mean-max</sub> | -0.003 ± 0.001 | 0.131 | **0.008** | -0.001 ± 0.001 | 0.186 | 0.550 | -0.002 ± 0.001 | 0.187 | **0.016** | 0.097 |

Where: CC-IMT<sub>mean</sub>, average of means of intima-media thickness in left and right common carotid arteries; CC-IMT<sub>max</sub>, the highest value of maximum intima-media thickness in left and right common carotid arteries; CC-IMT<sub>mean-max</sub>, mean of maximum intima-media thickness in left and right common carotid arteries. IMPROVE analyses were adjusted for (sex), age, population structure (MDS1-3), and use of lipid-lowering medication. UKB analyses were adjusted for (sex), age, population structure (PGC1-8), genotyping chip and use of lipid-lowering medication. $p < 0.05$ for statistical significance are highlighted in bold.

### Associations of PCSK9 variants with CC-IMT variables (mean, max and mean-max) in UKB

In UKB, a significant association was observed between rs11591147-T and CC-IMT_mean_, CC-IMT_max_ and CC- IMT_mean-max_ in sex-combined analyse and in men (S Table 4). The effect direction of these associations was consistent with that expected (i.e., beta <0) for CC-IMT_mean_ and CC-IMT_mean-max_ in sex-combined analyses, and for all CC-IMT variables in men. No other significant associations were observed; however the direction of the relationship was consistent with that expected for all CC-IMT variables in sex-combined analyses, and for CC-IMT_mean_ and CC-IMT_mean-max_ in men.

In UKB, analyses stratified by lipid-lowering medication demonstrated significant associations between rs11591147-T and CC-IMT variables in all participants, and between rs11591147-T and CC-IMT_max_ and CC- IMT_mean-max_ in untreated participants (S Table 5). The effect direction was as expected (beta <0) for CC- IMT_mean_-_max_ and CC-IMT_mean_-_max_ in all participants, and for CC-IMT_max_ and CC-IMT_mean_-_max_ in untreated participants.

### Associations of the lipid-lowering polygenic risk score (PRS) with CC-IMT in IMPROVE

In IMPROVE, the lipid-lowering PRS (combined effects of four genetic variants) was significantly associated (*p*<0.05) with all CC-IMT variables in women but not in men or the sex-combined analyses (Table 3). The PRS explained the 0.5% of the variance for CC-IMT_mean_ and CC-IMT_max_ (R^2^=0.005), and the 0.6% of the variance for CC-IMT_mean-max_ (R^2^=0.006). Beta values were in the expected direction (beta < 0) for all CC-IMT variables in the sex-combined analyses and in women, i.e., the higher the PRS, the more lipid-lowering alleles, the lower the values of CC-IMT_mean,_ CC-IMT_max_ and CC-IMT_mean-max_. The interaction between PRS and sex for CC-IMT_max_ was borderline statistically significant (*p*=0.051).

In IMPROVE, the analysis stratified by lipid-lowering medication demonstrated no significant associations between the PRS and CC-IMT variables (S Table 6). However, beta values were in the expected direction (beta < 0) for all CC-IMT variables in untreated subjects, and only in CC-IMT_mean_ and CC-IMT_mean-max_ in treated subjects. No significant PRS by lipid-lowering medication interaction was observed (S Table 6).

### Associations of the lipid-lowering polygenic risk score (PRS) with CC-IMT variables in UKB

In UKB, significant associations were observed between the PRS and CC-IMT variables in the sex-combined analyses and in men but not women (Table 3). In sex-combined analyses, the PRS explained approximately 20% of the variance for CC-IMT_mean_ (R^2^=0.193), CC-IMT_mean-max_ (R^2^=0.187) and the 14.5% for CC-IMT_max_ (R^2^=0.145). All beta values were in the expected direction (beta < 0). No significant interaction was observed between the PRS and sex in UKB (Table 3).

In UKB, analyses stratified by lipid-lowering medication in UKB (S Table 6) demonstrated significant associations between the PRS and all IMT measures in untreated individuals (all beta < 0), consistent with the findings observed in the entire group (S Table 6). The PRS explained approximately 20% of the variation in CC-IMT_mean_ and in CC-IMT_mean-max_, as well as about 14% of the variation in CC-IMT_max_ in untreated subjects (S Table 6). No significant interaction between the PRS and treatment was observed (S Table 6).

### Combined effects of PRS on CC-IMT variables in IMPROVE and UKB

Forest plots of PRS effects demonstrated a consistent negative impact on all C-IMT variables in IMPROVE, UKB and the meta-analyses (Figure 1), however the effect was more convincing for UKB (N=40512) and the meta-analyses (N=43404) than for IMPROVE (N=2892).

### PCSK9 levels in the IMPROVE study and association with the lipid-lowering PRS

Figure 2 shows the association between PRS and (adjusted geometric mean) PCSK9 levels in men and women (Panel A) as well as in treated and untreated individuals (Panel B). These PCSK9 levels were significantly higher in women compared to men (p<0.0001) and in treated compared to untreated individuals (p<0.0001).

**Figure 2.**
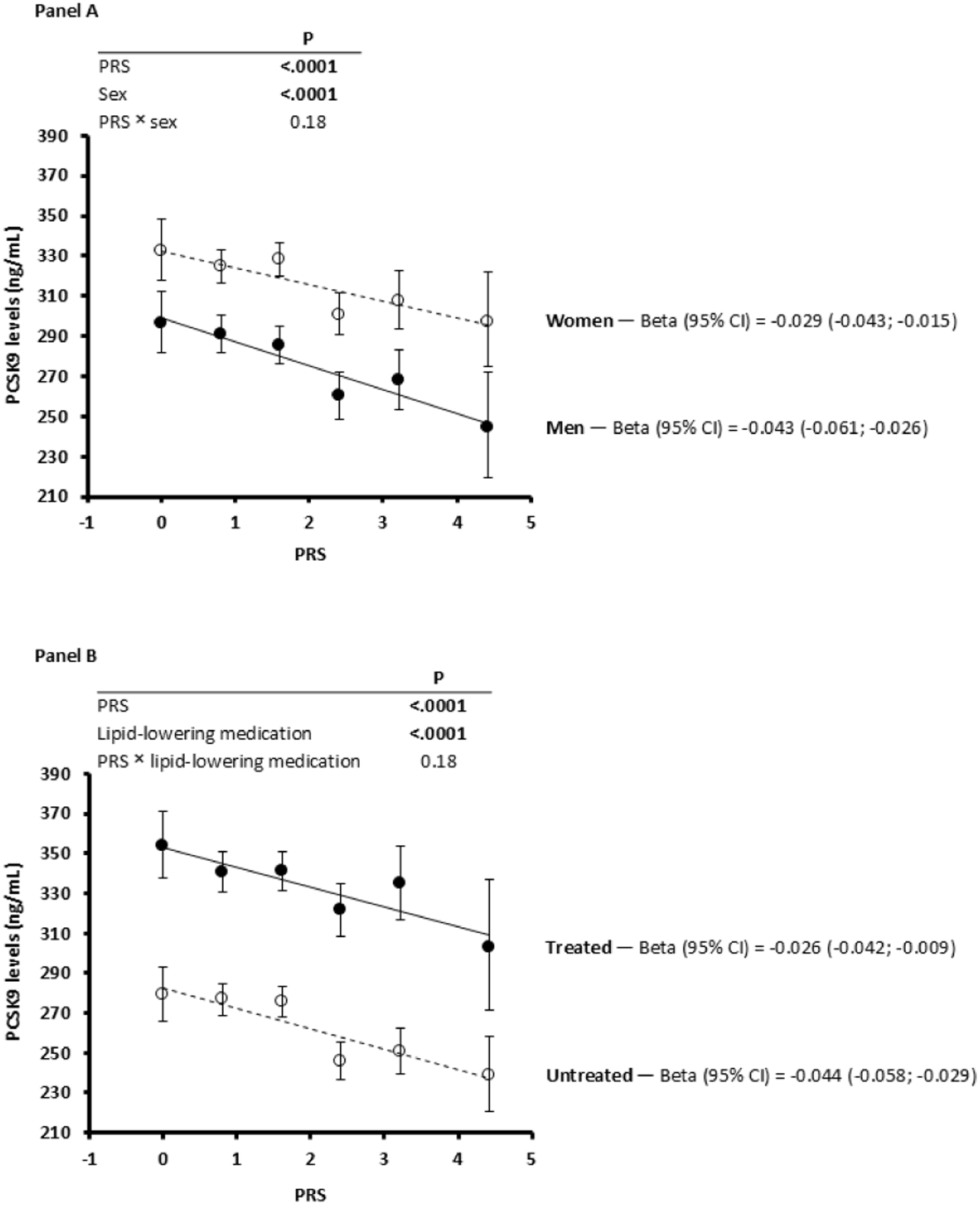
PCSK9 levels (adjusted geometric mean) Vs. PRS after participants’ stratification according to sex (Panel A) or use of lipid-lowering medication (Panel B) in the IMPROVE study. Beta (95% CI) refers to PCSK9 (natural log). The PRS was standardised prior to analysis. In Panel A, the analysis was adjusted for age, population structure (MDS1-3), and use of lipid-lowering medication. In Panel B, the analysis was adjusted for sex, age, and population structure (MDS1-3). The P_interaction_ was not statistically significant (i.e., P_interaction_ value < 0.05).

PCSK9 levels decreased with increasing PRS values and with no difference between men and women (P ∈ sex interaction=0.18) or between treated and untreated individuals (P ∈ lipid-lowering medication interaction=0.18). PCSK9 levels have not been measured in UKB.

## Discussion

Our results demonstrated that four genetic variants in the *PCSK9* locus, combined in a lipid-lowering PRS, were significantly associated with reduced atherosclerosis, assessed by C-IMT ultrasound measurements, in the high-CVD risk IMPROVE cohort (N=2892) and the general population UKB cohort (N=40512). The effect was modest and did not differ significantly by sex or use of lipid-lowering medication.

Genetic variants in the PCSK9 coding gene are not unique in influencing lipid and lipoprotein levels^38^, but they are among those with the greatest effect sizes. This is particularly true for the R46L variant (rs11591147-T), which has been associated with reduced PCSK9 levels ^39^, lower LDL-C levels ^19^, and a lower prevalence and/or incidence of ischemic heart disease ^40^. With the exception of the study by Mostaza et al., where no difference in LDL-C levels between carriers and non-carriers of the R46L variant was observed^17^, the negative association between this variant and LDL-C levels (well described in ad hoc meta-analysis^41^) has been observed in subjects from a variety of studies in various geographic regions, including the United States^19^, Canada^42^, the United Kingdom^43^, Australia^22^, Spain^20^, Denmark^21^, France^44^, Scotland^18^, Ireland^18^, and the Netherlands^18^.

This work extends the field by: firstly, demonstrating that the minor alleles of three *PCSK9* variants known to be associated with lower LDL-C levels (with MAFs of 2-17%) and the single *PCSK9* variant known to be associated with higher LDL-C levels (MAF=34%) were associated (albeit not all significantly) with circulating PCSK9 and LDL-C levels in IMPROVE. Secondly, we demonstrated that the variant with the strongest effect, rs11591147, was associated with C-IMT in UKB. That this association was not observed in IMPROVE is likely due to the smaller sample size. It is also worth noting that IMPROVE is a high CVD-risk cohort, with no prior CVD events, whereas the UKB is a general population cohort, which may include prior events. Thirdly, we demonstrated that the PRS (combined effects of the four variants) was associated with C-IMT measures in UKB, and when UKB and IMPROVE were meta-analysed. Similarly, no lipid-lowering medication interaction was observed, however it should be noted that the proportion of participants on lipid-lowering medication was vastly different between the two studies (49% in IMPROVE vs 12% in UKB). While cIMT is widely used as a marker of subclinical atherosclerosis ^2^ and is associated with increased CVD risk ^30^, it also reflects vascular remodeling processes such as endothelial hyperplasia and medial hypertrophy, which may be influenced by hemodynamic factors beyond lipid accumulation. The observed associations between *PCSK9* variants and cIMT underscore the role of LDL-C metabolism in arterial wall changes, supporting the potential utility of *PCSK9* genetic risk profiling in refining CVD risk prediction and early intervention strategies.

Our finding that these genetic variants and their PRS influence PCSK9 levels, LDL-C levels and C-IMT measures in a consistent fashion, is in line with the known role of PCSK9 on LDL-C levels and the impact of LDL-C on atherosclerotic burden. Recent work also suggests that *PCSK9* variants associated with lower rates of CVD were also associated with increased risk of respiratory diseases, including asthma, upper respiratory tract infections, and chronic obstructive pulmonary disease ^45^. As with most genetic effects on complex diseases, the magnitude of effect was small, even in the large UKB cohort. To our knowledge, this study is the first to provide evidence of a significant association, albeit modest, between carotid IMT and a PRS of variants associated with lipid levels by PCSK9. Indeed, the only studies available to date have investigated the relationship between lipid trait PRS associated with PCSK9 variants and clinical atherosclerosis (coronary ^46^ or peripheral ^47^), but not with subclinical atherosclerosis as is assessed here.

Sex differences in carotid IMT measurements have been described in previous studies ^30^. In IMPROVE, the PRS had stronger effects in women compared to men, with a borderline significant formal interaction test (*p*_interaction_=0.051). However, in UKB, the stronger effect was observed in men and the PRS x sex interaction test is null. Even if there is a sex effect of genetic variants in the *PCSK9* locus, large clinical trials (n∼500- 28000) of PCSK9 inhibitors have demonstrated no sex-difference in efficacy ^48, 49^.

### Strengths and limitations

This study benefits from several notable strengths. Firstly, the inclusion of two substantially different cohorts, IMPROVE (high CVD-risk, Pan-European recruitment) and the UKB (general population, UK recruitment), enhances the robustness and generalisability of the findings, particularly regarding the associations between *PCSK9* variants and carotid IMT. Secondly, the analyses focused on well-characterized genetic variants in the *PCSK9* gene with known functional effects, allowing for a robust examination of their impact on LDL-C levels and carotid artery atherosclerosis. Additionally, the use of a PRS to assess cumulative genetic effects provided a more comprehensive picture (as individual variants do not act in isolation). However, this study also had limitations that should be acknowledged. While significant associations were identified with the PRS, individual PCSK9 variants did not consistently demonstrate robust effects on carotid IMT, limiting the ability to draw firm conclusions about their independent contributions. Furthermore, the lack of PCSK9 level measurements in UKB restricts the capacity to fully correlate genetic variants with phenotypic outcomes, potentially obscuring the understanding of the underlying biological mechanisms. Finally, although the analysis adjusted for several known confounders, unmeasured factors—such as dietary habits, physical activity, or other genetic influences—may still affect the associations between PCSK9 variants and carotid IMT, introducing the possibility of bias in the results. In addition, UKB is well recognised and over-representative of the healthier parts of the general population^50^. A further consideration is that the analyses of UKB relied upon self-report of medication, which is well recognised as being impacted by recall bias. Overall, it is likely that biases in the data would result in underestimation (not overestimation) of effects.

## Conclusion

In conclusion, we have demonstrated genetic variants in the *PCSK9* locus which influence PCSK9 levels, and consequently LDL-C levels, also influences C-IMT and therefore is likely to be part of a causal pathway for atherosclerosis. Genetic risk profiling including these variants could be useful in future precision medicine strategies, where individuals most at risk can be targeted for intervention prior to overt disease development.

## Data Availability

UKB data is available via application to UKB (https://www.ukbiobank.ac.uk/). IMPROVE is not available for sharing. Analytic code is available upon request from the corresponding author.

## Author Contributions

Conceptualization, R.J. Strawbridge; methodology, R.J. Strawbridge, F. Veglia validation, R.J. Strawbridge, D. Coggi, N. Capra and F. Veglia; formal analysis, R.J. Strawbridge, D. Coggi, N. Capra and F. Veglia; investigation, M. Amato, B. Frigerio, A. Ravani, D. Sansaro, M.G. Lupo and C. Macchi; resources, R.J. Strawbridge, K. Savonen, M. Pirro, B. Gigante, and E. Tremoli; D. Baldassarre; N. Ferri data curation, R.J. Strawbridge, D. Coggi, J. Ward, N. Capra, M. Amato and D. Baldassarre; writing—original draft preparation, R.J. Strawbridge, and D. Coggi; writing—review and editing, D.M. Lyall, J. Ward, N. Ferri, M. Ruscica, A. Gallo, D.J. Mulder, R. Baetta, F. Veglia, J.P. Pell, P. Welsh, N. Sattar and D. Baldassarre; visualization, R.J. Strawbridge, D. Coggi and N. Capra; supervision, R.J. Strawbridge, and D. Baldassarre;project administration, R.J. Strawbridge, and D. Baldassarre; funding acquisition, E. Tremoli and D. Baldassarre.

All authors have read and agreed to the published version of the manuscript.

## Acknowledgements

We thank all participants and staff of the IMPROVE and UKB studies. This work uses data (UKB) provided by patients and collected by the NHS as part of their care and support.

